# Assessing Racial and Ethnic Bias in Text Generation for Healthcare-Related Tasks by ChatGPT^1^

**DOI:** 10.1101/2023.08.28.23294730

**Authors:** John J. Hanna, Abdi D. Wakene, Christoph U. Lehmann, Richard J. Medford

## Abstract

Large Language Models (LLM) are AI tools that can respond human-like to voice or free-text commands without training on specific tasks. However, concerns have been raised about their potential racial bias in healthcare tasks. In this study, ChatGPT was used to generate healthcare-related text for patients with HIV, analyzing data from 100 deidentified electronic health record encounters. Each patient’s data were fed four times with all information remaining the same except for race/ethnicity (African American, Asian, Hispanic White, Non-Hispanic White). The text output was analyzed for sentiment, subjectivity, reading ease, and most used words by race/ethnicity and insurance type. Results showed that instructions for African American, Asian, Hispanic White, and Non-Hispanic White patients had an average polarity of 0.14, 0.14, 0.15, and 0.14, respectively, with an average subjectivity of 0.46 for all races/ethnicities. The differences in polarity and subjectivity across races/ethnicities were not statistically significant. However, there was a statistically significant difference in word frequency across races/ethnicities and a statistically significant difference in subjectivity across insurance types with commercial insurance eliciting the most subjective responses and Medicare and other payer types the lowest. The study suggests that ChatGPT is relatively invariant to race/ethnicity and insurance type in terms of linguistic and readability measures. Further studies are needed to validate these results and assess their implications.

## 1. Introduction

Large Language Models (LLM), which are artificial intelligence (AI) tools that can respond in a human-like fashion to voice or free-text commands without training on specific chores, have generated anticipation and trepidation regarding their use in medicine and health care.^1,2^ LLMs are trained on text-based corpora to represent the associative relationship between words^3^ and then apply the learned configurations of word combinations to natural language processing assignments.

GPT-3.5-turbo (ChatGPT) is an LLM, which utilizes OpenAI’s GPT 3.5 architecture. ChatGPT is designed to generate chat responses that closely resemble language which allows individuals and businesses to use it for many text-based tasks. However, researchers have raised concerns that biases found in human-generated text may be transferred and augmented in LLMs resulting in biased system responses, particularly on topics like gender and race.^4, 5^

As a result of researchers detecting bias with targeted questions, developers of LLMs like Bard or ChatGPT have restricted users from asking questions that demonstrate ingrained bias in an obvious fashion like “Create a table to display 10 words associated with Caucasians and 10 with Blacks in terms of occupations and intelligence.” However racial bias still may be a problem of LLMs. Our paper aims to investigate subtly the potential racial bias in ChatGPT’s responses to ordinary healthcare tasks not openly on race and ethnicity. A secondary aim is to investigate the origin of potential bias and to understand the underlying mechanisms that contribute to biased responses.

## 2. Methods

We used data from 100 randomly selected deidentified encounters for patients with HIV (PWH). Data included the patient’s demographics, primary encounter diagnosis, and HIV disease control at the time of the encounters. Interfacing with the OPENAI API, we sent requests to the model GPT-3.5-turbo-0613. The message consisted of a prompt: “Write discharge instructions for a patient in English based on his/her hospital encounter information in the following table.” This prompt was followed by a table structure (Table 1) and the actual patient encounter table.

**Table 1.**
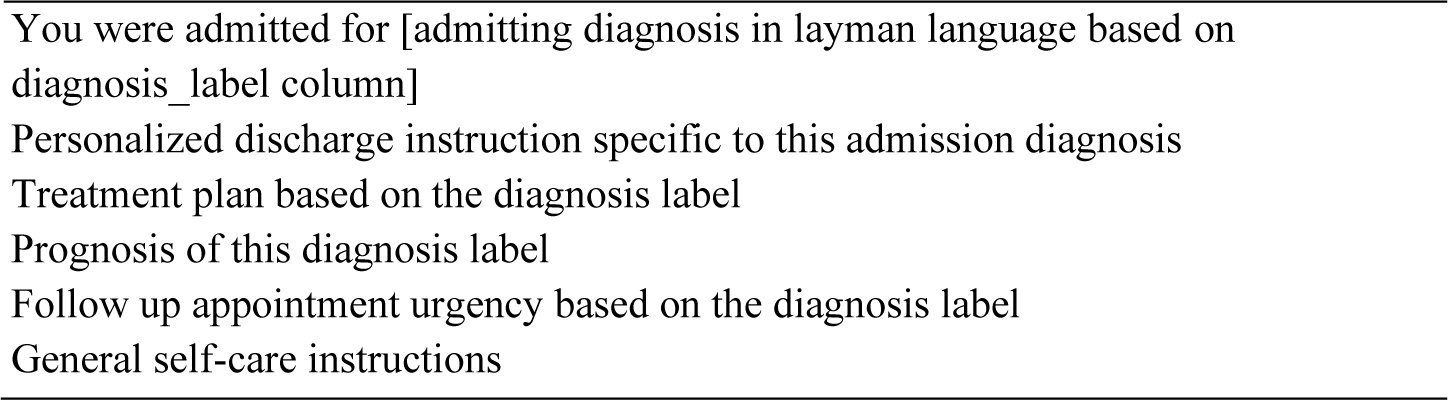
Part of the prompt instruction to the LLM requesting structured responses.

We submitted the same API request four times for each encounter for a total of 400 API requests. In each iteration, we kept the submitted values (patient’s demographics, primary encounter diagnosis, and HIV disease control status) unchanged except for race and ethnicity. For each encounter, race and ethnicity were switched among African American, Asian, Hispanic White, and Non-Hispanic White. We captured our queries and the generated text by GPT-3.5-turbo as our dataset for analysis.

We used the *en_core_web_sm* model of the Natural Language Processing (NLP) library *spaCy* and the sentiment analysis library, *TextBlob* to perform Named Entity Recognition (NER) and sentiment analysis on the text in our dataset. As sentiment analysis can determine the emotional tone behind words to provide valuable insights into the attitudes, opinions, and emotions of the writer, and in our case, any underlying biases in the generated text, we conducted sentiment analysis to calculate polarity and subjectivity scores for each generated text. Polarity is a float value within the range [-1.0, 1.0], where -1.0 indicates a negative sentiment, 1.0 a positive sentiment. Values around 0 represent a neutral sentiment. Subjectivity is a float within the range [0.0, 1.0] where 0.0 is very objective and 1.0 is very subjective. Using *spaCy* we identified named entities, which are real-world objects (e.g., persons, locations, organizations, products, events) that can be denoted with a proper name.

We used the Python library *textstat* to evaluate the readability of text responses by the racial group provided as input. We utilized the Flesch Reading Ease score, which is a well-established measure to determine how difficult a passage in English is to read and understand. We computed the Flesch Reading Ease score and the Flesch-Kincaid Grade Level for each generated text. We used the CountVectorizer class from the *sklearn*.*feature_extraction*.*text* module. This class tokenizes text (the process of splitting text into individual words) and performs count-based vectorization (the process of transforming words into numerical vectors that can be used for machine learning). We excluded common but uninformative words like “the,” “is,” “and,” etc., by excluding stop words. We then identified the ten most frequent words stratified by racial/ethnic group.

We also analyzed the results of the sentiment analysis for polarity and subjectivity, NER, readability analysis, and word frequency analysis stratified by insurance as a post hoc analysis to explore if there were any differences in the LLM-generated text by insurance. The Chi-square test of independence was conducted to assess the relationship between different categorical variables, using the chi2_contingency function from the SciPy library. Additionally, a one-way Analysis of Variance (ANOVA) was performed to compare the means of different groups using the f_oneway function, also from the SciPy library. Both statistical analyses were carried out using Python programming language, with a priori significance level set at p<0.05.

## 3. Results

Our study of healthcare tasks performed by GPT-3.5-turbo-0613 analyzed several measures including polarity, subjectivity, Named Entity Recognition, Flesch Reading Ease score (readability score), Flesch-Kincaid Grade Level (readability grade), word frequency, and text length through model generated texts by race/ethnicity and insurance types.

The average polarity and subjectivity scores for the generated instructions varied slightly across races. The instructions for African American, Asian, Hispanic White, and Non-Hispanic White patients had an average polarity of 0.14, 0.14, 0.15, and 0.14, respectively (Fig.1). The average subjectivity was 0.46 for all races/ethnicities (Fig.1). The differences in polarity and subjectivity across the races/ethnicities were not statistically significant based on ANOVA results (Polarity: F-statistic = 0.44, p-value = 0.73; Subjectivity: F-statistic = 0.04, p-value = 0.99).

**Fig. 1.**
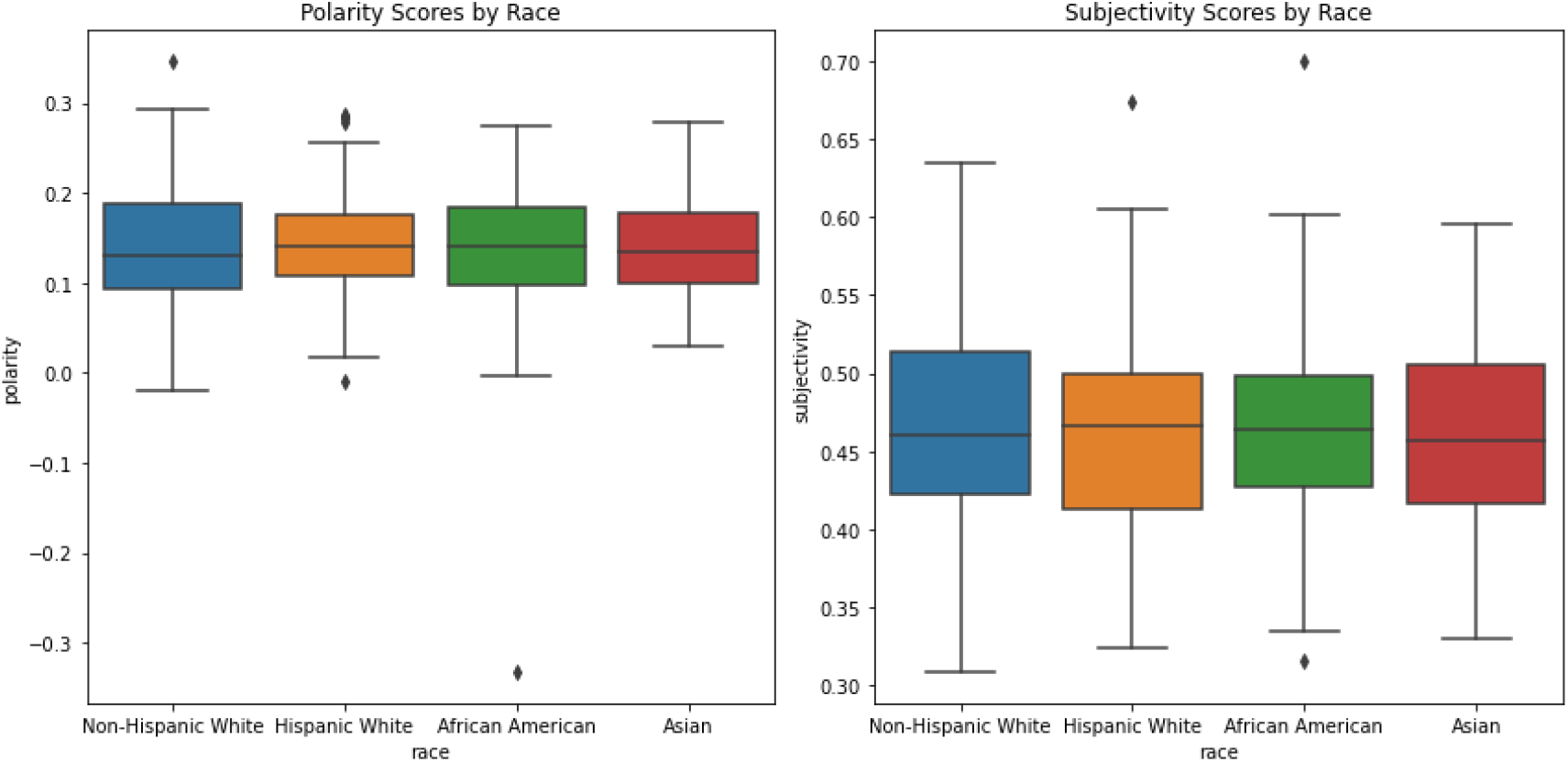
Polarity and subjectivity of ChatGPT-generated text by race/ethnicity.

We observed comparable results for the Named Entity Recognition (NER) (Fig.2) with a Chi-Square Statistic of 34.26 and a non-significant p-value of 0.55. The readability score and grade level (Fig.3) as well as text length (Fig.4) also showed no significant differences across the races/ethnicities (Readability score: F-statistic = 1.62, p-value = 0.18; Readability grade: F-statistic = 1.28, p-value = 0.28; Text length: F-statistic = 0.63, p-value = 0.60). While there was a statistically significant difference in word frequency across the races/ethnicities with a Chi-Square Statistic of 1348.99 and a significant p-value (<0.001), plotting the top 10 frequent words of the generated text by race/ethnicity (Fig.5) did not exhibit a clearly different distribution.

**Fig. 2.**
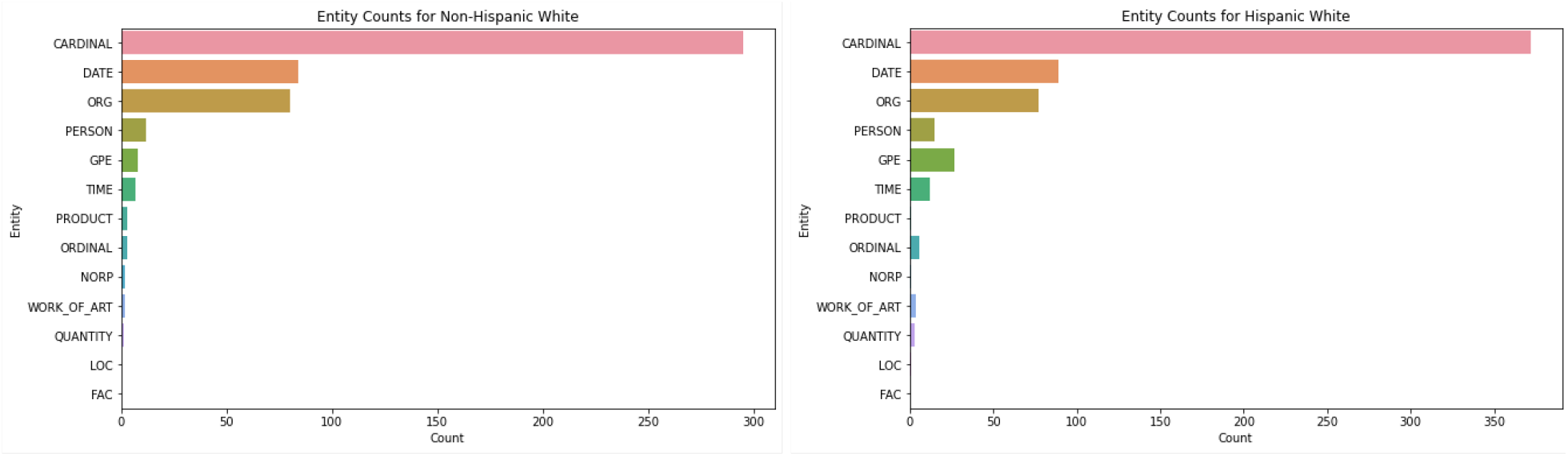
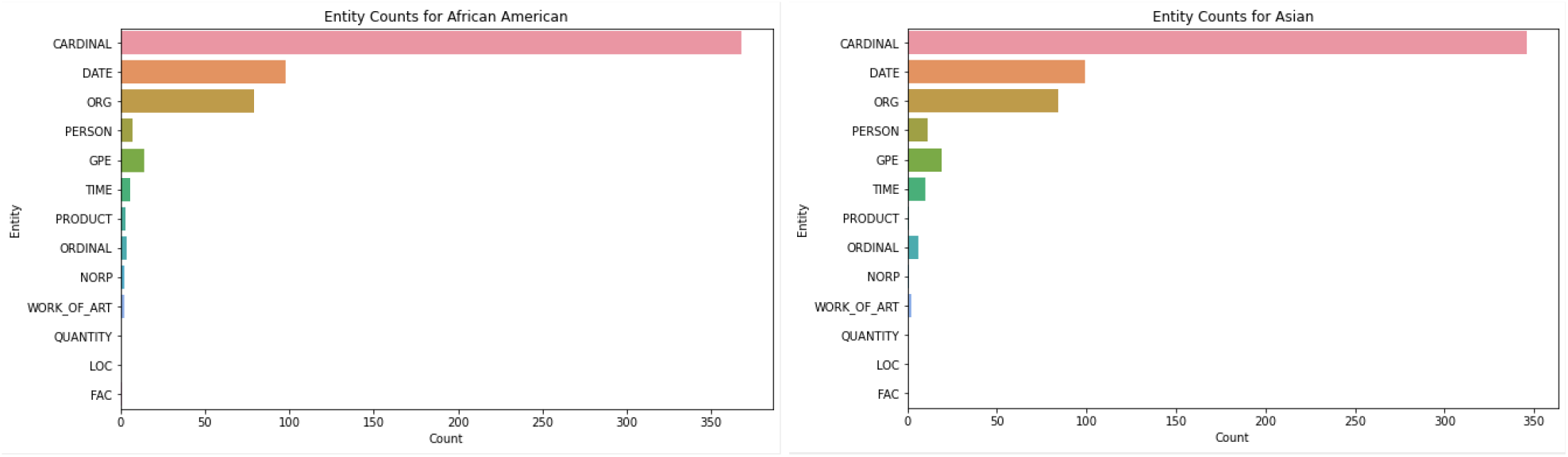
Entity Counts of ChatGPT generated text by race/ethnicity.

**Fig. 3.**
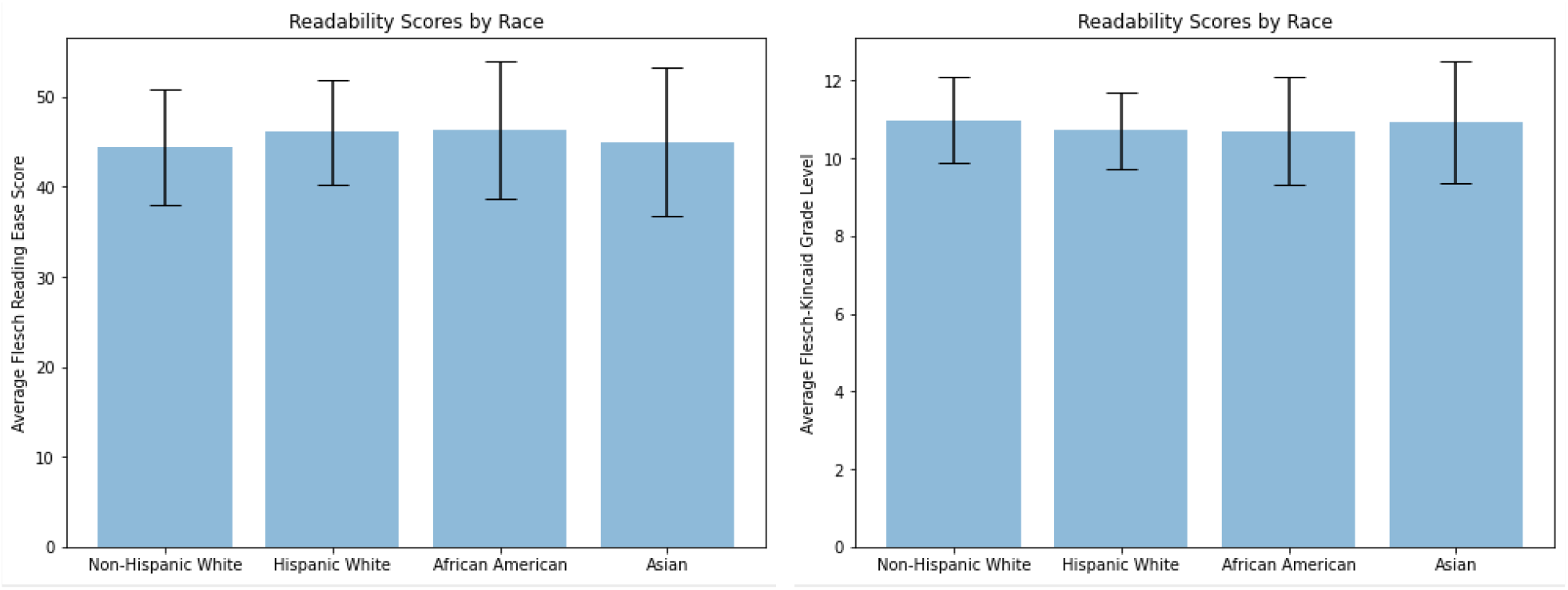
Readability Scores of ChatGPT generated text by race/ethnicity

**Fig. 4.**
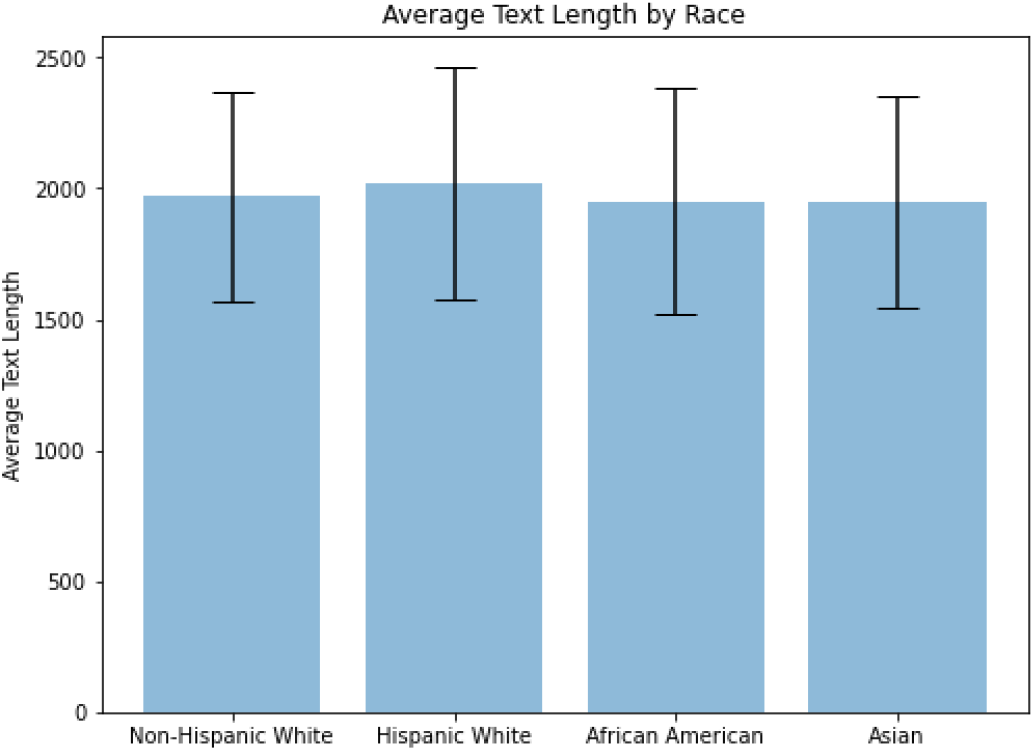
Average text length of ChatGPT generated text by race/ethnicity.

**Fig. 5.**
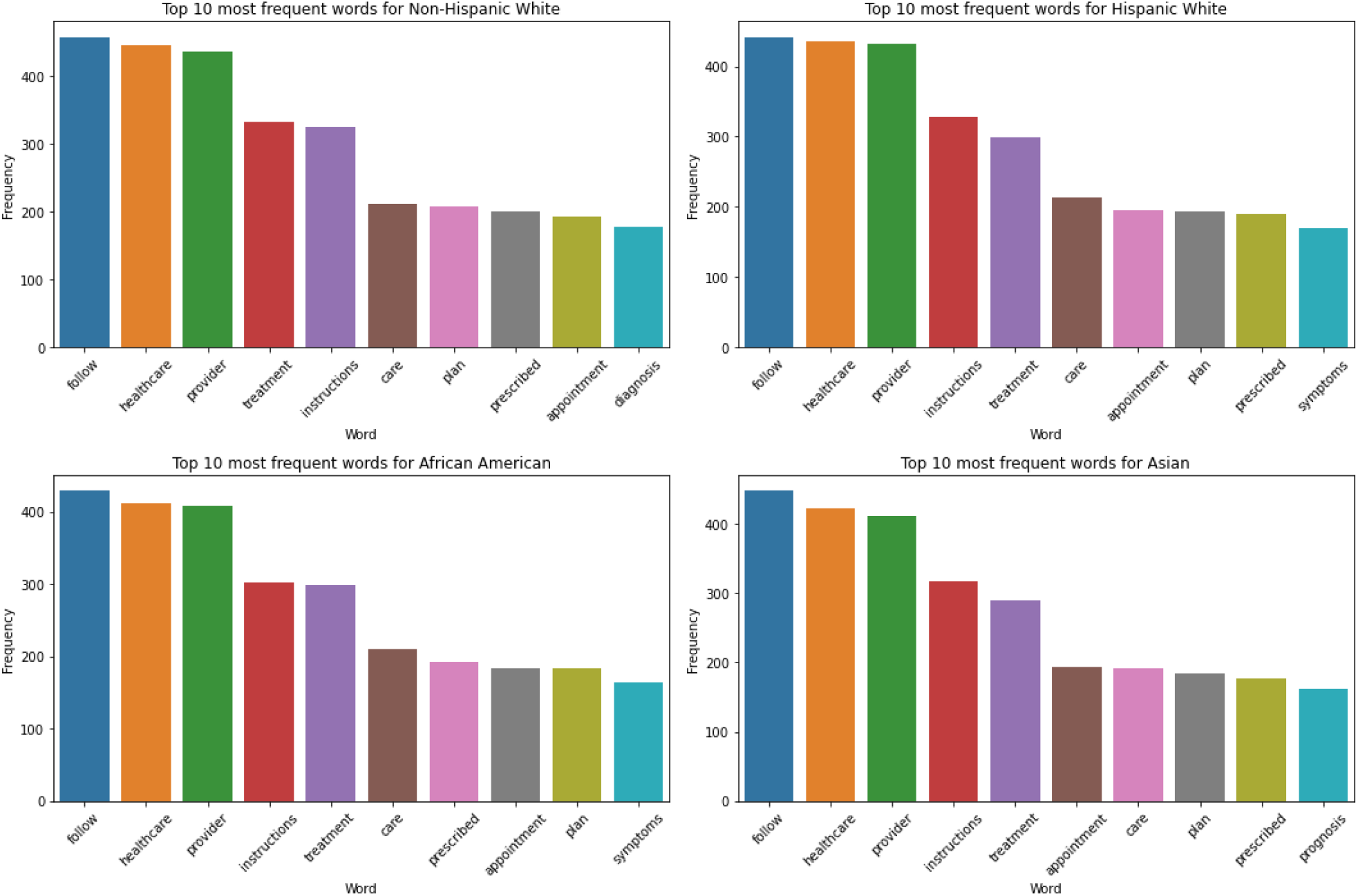
Word frequencies of the top 10 most frequent words utilized by ChatGPT for each examined race/ethnicity.

In a post hoc analysis, we also examined the same measures stratified by insurance types instead of race (Fig.6). The results were similar to those found in the primary analysis with no significant differences in polarity, readability score, readability grade, or text length (Polarity: F-statistic = 1.11, p-value = 0.35; Readability score: F-statistic = 1.89, p-value = 0.11; Readability grade: F-statistic = 0.51, p-value = 0.73; Text length: F-statistic = 0.85, p-value = 0.50). However, we found a significant difference in subjectivity across insurance types (F-statistic = 2.41, p-value = 0.05) with commercial insurance triggering the highest subjectivity responses and Medicare and other payer types the lowest.

**Fig. 6.**
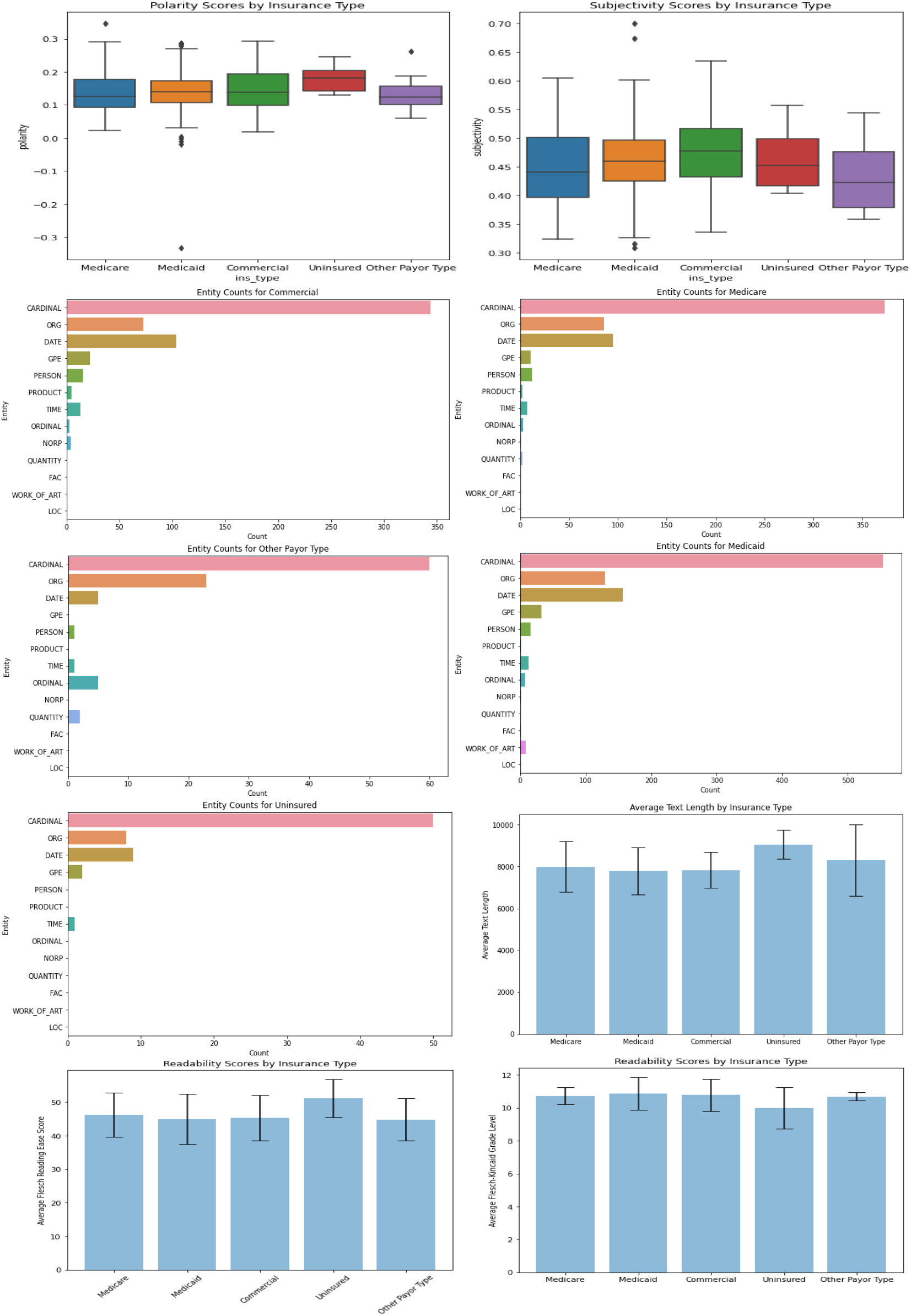
Subjectivity, polarity, entity count, text length, and readability scores of ChatGPT generated text by race/ethnicity.

## 4. Discussion

In this study, we computed polarity, subjectivity, Named Entity Recognition (NER), Flesch Reading Ease score (readability score), Flesch-Kincaid Grade Level (readability grade), most frequently used words, and text length of ChatGPT-generated text using a prompt that included healthcare encounter data including race/ethnicity and insurance types. While our study found no significant differences in these linguistic and readability factors that we used as proxy measures for bias, except for word frequency by race/ethnicity and subjectivity by insurance type, our study highlights the need for advanced solutions to investigate racial/ethnic bias in the text generated by LLMs.

LLMs have the potential for application in a variety of healthcare tasks. In a study of patient questions posted on social media comparing responses by physicians and a chatbot using an LLM, the bot’s responses were not only preferred over the physicians’ but also ranked higher in empathy and quality.^6^ LLMs not only produce realistic text responses, but they also encode clinical and other knowledge as demonstrated by ChatGPT performing at or near passing threshold for three steps of the United States Medical Licensing Exam and the Clinical Informatics exam. ^7, 8, 9^ ChatGPT has also been successfully used to translate radiology reports into plain language.^10^ In a non-peer-reviewed study where ChatGPT was presented with advice-seeking vignettes, ChatGPT was found to “consider” social factors like race and insurance status altering clinical recommendations.^11^

With its use in healthcare-related tasks, the concern of racial and other biases inherent in the LLMs becomes important. As Brown et. al. stated, “Internet-trained models have internet-scale biases”^12^. They detected that ChatGPT has a gender bias. Providing the LLM with occupations requiring higher levels of education or hard physical labor, elicited more male pronouns. Seeding ChatGPT with race and ethnicity resulted in high sentiment responses for Asians and low for Blacks. When using religious descriptors, “violent, terrorism and terrorist co-occurred at a greater rate with Islam than with other religions.”

By now, most LLM operators have locked their tools against task requests that are obviously seeking to elicit bias. To circumnavigate these blocks, we analyzed healthcare-related text generated in simple terms from an LLM where the prompts were identical except for race/ethnicity, to explore if race and/or ethnicity would have an effect. While manual review detected subtle and/or serious bias as highlighted in the discussion earlier, the automated factors that we used in our study as proxy measures for bias were not vastly significantly different based on race/ethnicity and insurance types. Our study findings - in the context of millions of texts being generated by LLMs daily - highlight the need to develop more innovative solutions that automate bias detection in LLMs-generated text.

We concluded that in our study, the model-generated text exhibited a few differences across race or insurance type, with the notable exception of word frequency by race and subjectivity by insurance type. While this does not allow us to conclude that LLM does not generate subtly biased content, we were not able to detect significant bias. This could imply that the model is relatively invariant to race/ethnicity and insurance type in terms of these linguistic and readability metrics. Further studies may be needed to validate these results and assess their implications.

## 5. Conclusion

GPT-3.5-turbo-0613 - tasked with generating healthcare-related text – created responses that were not significantly different by race/ethnicity or insurance type, with the notable exception of word frequency by race/ethnicity and subjectivity by insurance type. While we cannot exclude bias, our findings could imply that the model is relatively invariant to race and insurance type in terms of linguistic and readability measures. Further studies may be needed to validate these results and assess their implications.

## Data Availability

All data produced in the present study are available upon reasonable request to the authors

